# Gout, rheumatoid arthritis and the risk of death from COVID-19: an analysis of the UK Biobank

**DOI:** 10.1101/2020.11.06.20227405

**Authors:** Ruth K Topless, Amanda Phipps-Green, Megan Leask, Nicola Dalbeth, Lisa K Stamp, Philip C Robinson, Tony R Merriman

## Abstract

**Objectives:** To assess whether gout and / or rheumatoid arthritis (RA) are risk factors for coronavirus disease 19 (COVID-19) diagnosis. To assess whether gout and / or RA are risk factors for death with COVID-19.

**Methods:** We used data from the UK Biobank. Multivariable-adjusted logistic regression was employed in the following analyses: Analysis A, to test for association between gout or RA and COVID-19 diagnosis (n=473,139); Analysis B, to test for association between gout or RA and death with COVID-19 in a case-control cohort of people who died or survived with COVID-19 (n=2,059); Analysis C, to test for association with gout or RA and death with COVID-19 in the entire UK Biobank cohort (n=473,139)

**Results:** RA, but not gout, associated with COVID-19 diagnosis in analysis A. Neither RA nor gout associated with risk of death in the COVID-19-diagnosed group in analysis B. However RA associated with risk of death related to COVID-19 using the UK Biobank cohort in analysis C independent of comorbidities and other measured risk factors (OR=1.9 [95% CI 1.2 ; 3.0]). Gout was not associated with death related to COVID-19 in the same UK Biobank analysis (OR=1.2 [95% CI 0.8 ; 1.7]).

**Conclusion:** Rheumatoid arthritis is a risk factor for death with COVID-19 using the UK Biobank cohort. These findings require replication in larger data sets that also allow inclusion of a wider range of factors.

**Key messages:** Information on the risk of death from COVID-19 for people with gout and rheumatoid arthritis is scarce.

In an analysis of the UK Biobank there is an increased risk of death related to COVID-19 for people with rheumatoid arthritis independent of included co-morbidities, but not gout.

The findings need to be replicated in other datasets where the influence of therapies for rheumatoid arthritis can be tested.

## Background

Data on coronavirus disease 2019 (COVID-19) outcomes for people with the two most common inflammatory arthropathies, gout and rheumatoid arthritis (RA), are scarce. An international registry study of 600 people with rheumatic diseases did not report any data on association of gout with hospitalisation, owing to the small number of people with gout included (1). In the same study, people with RA did not have a different risk of hospitalisation compared to other rheumatic diseases (1). In the OpenSAFELY study (2) that compared risk factors for 10,926 people who died from COVID-19 to the general population in the UK, RA was pooled with systemic lupus erythematosus and psoriasis, this combined group had a hazard ratio of 1.2 [95% CI: 1.1;1.3] for death. However gout was not examined in the OpenSAFELY study. A population-based study in Denmark reported a hazard ratio of 1.4 [0.8 ; 2.5] for an outcome of mechanical ventilation or severe respiratory disease or death with COVID-19 in people with RA (3). In a US study comparing people with COVID-19 with systemic autoimmune rheumatic diseases, of whom 45% had RA, to people with COVID-19 without these diseases there was increased risk of hospitalisation and admission to intensive care but not increased risk of death (4). A Spanish study reported no evidence for association of chronic inflammatory arthritis (48% with RA) with poor outcome in people with COVID-19 (5).

Gout is caused by an exuberant auto-inflammatory interleukin-1β-driven innate immune system response to monosodium urate crystals (6). Theoretically this has the potential to lead to an increased immune response to severe acute respiratory syndrome coronavirus 2 (SARS-CoV-2). Poorer COVID-19 outcomes have been associated with high serum levels of IL-6, IL-8 and TNF-α (7), raising the possibility that people with gout might be at risk of a poor outcome due to the fact that they also have higher circulating levels of these factors (8). Gout is also strongly associated with cardiometabolic co-morbidities such as type 2 diabetes, kidney disease and heart disease (9), all established risk factors for COVID-19-related mortality (2). Gout medications may also influence outcomes following the development of COVID-19: two randomised control trials of colchicine, which is widely used as prophylaxis and treatment for the gout flare (10), reported better clinical outcomes including a shorter time in hospital and shorter duration requiring supplemental oxygen in people hospitalised with COVID-19 in those randomised to colchicine (11,12). There is also non-randomised evidence of the efficacy of colchicine in COVID-19 in a small case-control study (13).

Rheumatoid arthritis is a T-cell and B-cell mediated autoimmune disease that primarily affects the joints but also includes systemic manifestations. Like gout, RA is an independent risk factor for cardiovascular disease (14). The profile of RA includes increased levels of pro-inflammatory cytokines TNF-α and IL-6 (15), a similar profile to COVID-19 (16), with the potential to lead to an increased immune response to infection by SARS-CoV-2.

The aim of this study was to determine if gout and RA are risk factors for COVID-19 diagnosis or death with COVID-19.

## Participants and methods

### Data availability

This research was conducted using the UK Biobank Resource (approval number 12611). The UK Biobank is a large resource of nearly 500,000 volunteers aged 49-86 years of age at recruitment. Recruitment began 2006 with follow-up for at least 30-years (17). SARS-CoV-2 test information, ICD-10 hospital codes, death records and general practice prescription information were obtained via the UK Biobank data portal on the 16^th^ of September 2020. This information covered hospital diagnoses between 1991 and 30^th^ June 2020, SARS-CoV-2 tests between 16^th^ March and 24^th^ August 2020, and death records up until 14^th^ August 2020.

### Gout, RA and COVID-19 definitions and case-control datasets

The criteria for COVID-19 diagnosis was defined as participants with 1) a positive SARS-CoV-2 test and / or 2) ICD-10 code for confirmed COVID-19 (U07.1) or probable COVID-19 (U07.2) in hospital records, or death records (Figure 1). This definition resulted in identification of 2,118 individuals who were further divided into those that died (n=457) based on death records and those that were known to survive (n = 1,602). Fifty-nine participants who were diagnosed after 26^th^ July 2020 (28 days before the last recorded death) were removed from the cohort used in Analysis B (below) given the unknown outcome in these individuals. Gout was ascertained by a previously validated gout definition using the following criteria: self-reported gout (visits 0-2); taking allopurinol or sulphinpyrazone therapy either by self-report or from linked general practice scripts (excluding those who also had hospital diagnosed lymphoma or leukemia ICD10 C81 - C96); or hospital-diagnosed gout (ICD-10 code M10) (18), this case definition has been validated (18,19). The gout case-control cohort (n = 473,139) consists of 13,105 cases (gout) and 460,034 controls (non-gout). RA affection was ascertained using a combination of self-report of RA at more than one study visit, hospital recorded RA (ICD-10 code M05 - M06) on more than one occasion, or a self-report of RA at recruitment and at least one hospital record of RA. The RA cohort (n = 473,139) consists of 5,409 people with and 467,730 people without RA. For the RA and gout cohorts we developed three case-control datasets to test for association with the following outcomes:

**Figure 1.**
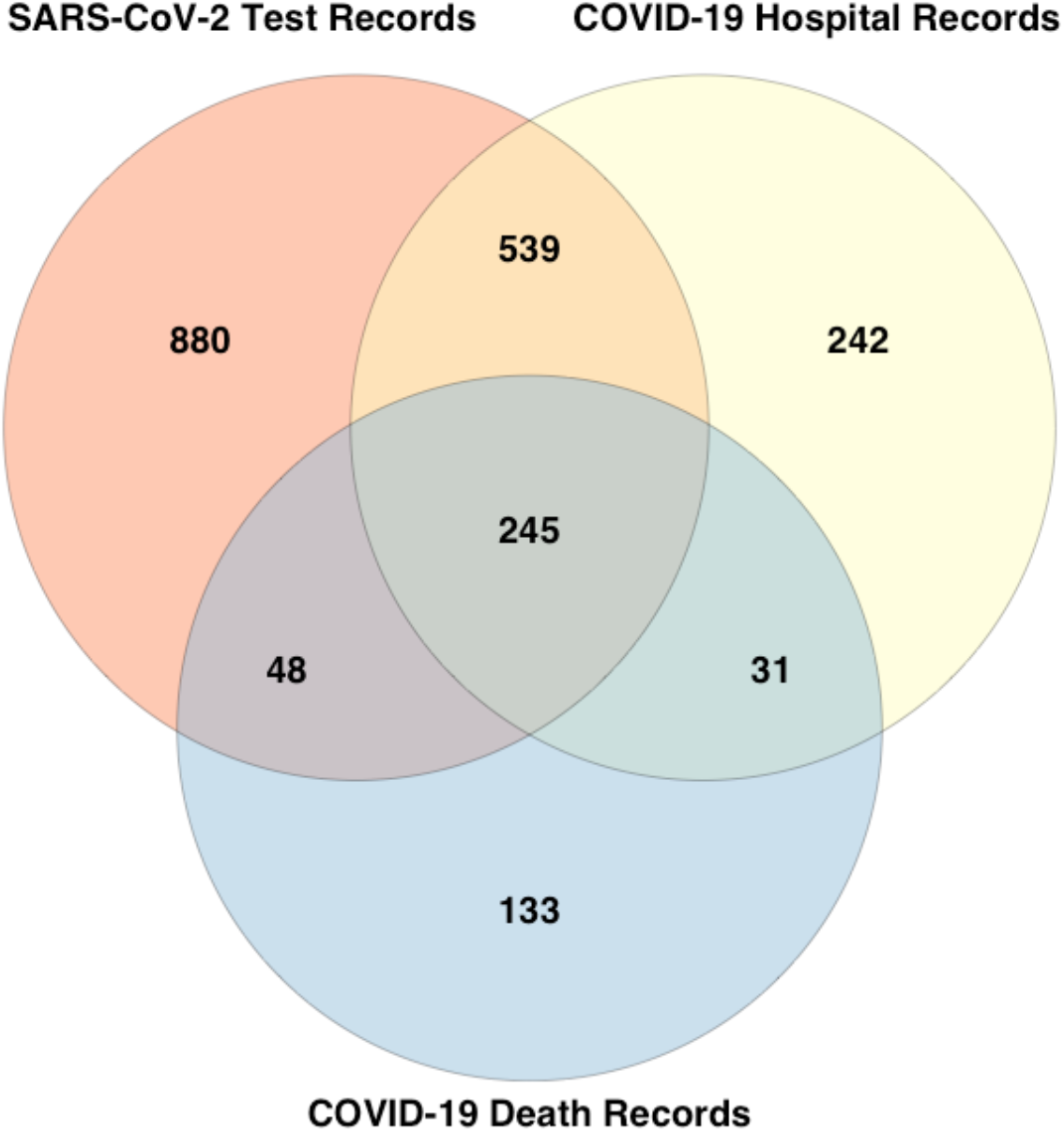
Data sources of COVID-19-diagnosed individuals. Of the 2,118 COVID-19-diagnosed individuals, 1,712 were identified from positive SARS-CoV-2 test results (880 unique to this group), 1057 identified from hospital records (242 unique to this group), and 457 identified from death records (133 unique to this group).

Dataset A (Analysis A) to test for association with COVID-19 diagnosis in a population-based cohort. There were 2,118 cases and 471,021 controls.

Dataset B (Analysis B) to test for association with death from COVID-19 in people with COVID-19. There were 457 people diagnosed with COVID-19 who died and 1,602 people diagnosed with COVID-19 who survived.

Dataset C (Analysis C) to test for association with death related to COVID-19 in a population-based cohort. There were 457 people diagnosed with COVID-19 who died and 472,682 others that included 1,616 people diagnosed with COVID-19 not known to have died.

### Ethnicity, age and comorbidity data

Self-reported ethnicity was grouped into White British (British, Irish, White, Any other white background), Black British (African, White and Black African, Black or Black British, Caribbean, White and Black Caribbean, Any other Black background), Asian British (Asian or Asian British, Chinese, Indian, Pakistani, Bangladeshi, White and Asian, Any other Asian background), and Other (Other ethnic group, Mixed, Any other mixed background, Do not know, Prefer not to answer). Age was calculated for 2020 from year of birth. The ICD-10 hospital codes used to determine additional comorbidity status were: C00 – C96 (cancer), D80 - D89 (immunodeficiencies), E08 - E13 (diabetes mellitus), E78 (disorders of lipoprotein metabolism and other lipidemias), F01-F03 (dementia), I10 - I15 (hypertensive diseases), I60 - I69 (cerebrovascular diseases), I20 - I25 (ischemic heart diseases), I26 - I28 (pulmonary heart disease), I50 (heart failure), J44 (chronic obstructive pulmonary diseases), J45 (asthma), M19.9 (osteoarthritis) and N18 (chronic kidney disease).

### Statistical analysis

All association analyses were done using R v4.0.2 in RStudio 1.2.5019. Age groups used in the analysis were <60 years (n= 89,607), 60-69 years (n= 151,139), 70-74 years (n=110,159) and >74 years (n= 122,222). Two models were used: adjustment with age group, sex, ethnicity, Townsend deprivation index, BMI, smoking status (Model 1); and Model 1 plus adjustment by the 15 other co-morbidities evaluated (Model 2). A *P* < 0.05 threshold indicated nominal evidence for association.

## Results

### Association with diagnosis of COVID-19

Results from the association analyses of gout and RA with COVID-19 diagnosis (Analysis A) using Model 1 (adjustment by current age, sex, Townsend deprivation index, ethnicity groups, body mass index and smoking status) are presented in Table 2. Both gout and RA associated with an increased risk of COVID-19 diagnosis of 1.5-fold [95% CI: 1.2 ; 1.8] and 2.2-fold [1.7 ; 2.9], respectively. We also included in our study other diseases known to be risk factors for poor COVID-19 outcome (2) both for comparison of effect sizes and inclusion in models as potential confounders. In comparison, data for cerebrovascular diseases were (OR 4.7 [4.1 ; 5.3]), heart failure (OR 4.0 [3.5 ; 4.7]), chronic kidney disease (OR 3.7 [3.3 ; 4.3]), pulmonary heart disease (OR 3.4 [2.8 ; 4.1]), immunodeficiencies (OR 3.3 [2.2 ; 4.7]) and chronic obstructive pulmonary disorders (OR 3.1 [2.7 ; 3.6]). Two to three-fold increases in risk were estimated for hypertensive diseases (OR 2.4 [2.2 ; 2.7]), diabetes mellitus (OR 2.4 [2.1 ; 2.7]), and lipoprotein disorders (OR 2.1 [1.9 ; 2.4]). One to two-fold increases in risk were estimated for ischemic heart diseases (OR 2.0 [1.7 ; 2.2]), cancer (OR 1.7 [1.6 ; 1.9]), asthma (OR 1.6 [1.4 ; 1.8]), osteoarthritis (OR 1.5 [1.4 ; 1.7]), and dementia strongly associated with COVID-19 (OR 18.2 [15.5 ; 21.4]). After adjustment by Model 1 variables and the additional 15 co-morbidities evaluated (Model 2), gout no longer associated with COVID-19 diagnosis, nor did ischemic heart disease, asthma and osteoarthritis (Table 3). RA maintained nominal association (OR 1.3 [1.0 ; 1.8]). Age was associated with decreased risk of COVID-19 diagnosis (OR 0.54 [0.47 ; 0.61], OR 0.45 [0.39 ; 0.53], OR 0.60 [0.52 ; 0.69] for 60-69 yrs, 70-74 yrs and >74 years, respectively, when compared to <60 yrs (Table 3). This decreased risk may reflect a number of factors that influence exposure to SARS-CoV-2 in this age group, including public health messaging around limiting exposure for older people.

**Table 1.** Characteristics of participants with and without gout and rheumatoid arthritis

|  | Gout | Non-Gout | Rheumatoid<br>Arthritis | Non-RA |
| --- | --- | --- | --- | --- |
| n | 13,105 | 460,034 | 5,409 | 467,730 |
| Age, <60 years (%) | 1,110 (8.5) | 88,499 (19.2) | 465 (8.6) | 89,144 (19.1) |
| Age, 60 - 69 years (%) | 3,285 (25.1) | 147,857 (32.1) | 1,466 (27.1) | 149,676 (32.0) |
| Age, 70 - 74 years (%) | 3,457 (26.4) | 106,705 (23.2) | 1,470 (27.2) | 108,692 (23.2) |
| Age, >74 years (%) | 5,253 (40.1) | 116,973 (25.4) | 2,008 (37.1) | 120,218 (25.7) |
| Males (%) | 11,253 (85.9) | 200,374 (43.6) | 1,537 (28.4) | 210,090 (44.9) |
| Females (%) | 1,852 (14.1) | 259,660 (56.4) | 3,872 (71.6) | 257,640 (55.1) |
| BMI, mean (SD) | 30.4 (5.0) | 27.3 (4.7) | 28.5 (5.6) | 27.4 (4.8) |
| Townsend deprivation index,<br>mean (SD) | -1.07 (3.2) | -1.34 (3.1) | -0.86 (3.3) | -1.34 (3.1) |
| White British ethnicity (%) | 12,321 (94.2) | 432,226 (94.1) | 5,042 (93.5) | 439,505 (94.1) |
| Black British ethnicity (%) | 368 (2.8) | 11,458 (2.5) | 161 (3.0) | 11,665 (2.5) |
| Asian British ethnicity (%) | 189 (1.5) | 8,599 (1.9) | 114 (2.1) | 8,674 (1.9) |
| Other ethnicities (%) | 196 (1.5) | 6,940 (1.5) | 77 (1.4) | 7,059 (1.5) |
| Never smoke (%) | 5,612 (43.2) | 256,879 (56.2) | 2,495 (46.6) | 259,996 (55.9) |
| Current smoker (%) | 6,202 (47.7) | 154,574 (33.8) | 2,210 (41.3) | 158,566 (34.1) |
| Ex-smoker (%) | 1,190 (9.2) | 46,005 (10.1) | 650 (12.1) | 46,545 (10.0) |
| Asthma (%) | 1,562 (11.9) | 39,336 (8.6) | 1,073 (19.8) | 39,825 (8.5) |
| Cancer (%) | 2,653 (20.2) | 64,197 (14.0) | 1,135 (21.0) | 65,715 (14.1) |
| Cerebrovascular diseases (%) | 939 (7.2) | 13,154 (2.9) | 410 (7.6) | 13,683 (2.9) |
| Chronic kidney disease (%) | 1,844 (14.1) | 11,489 (2.5) | 570 (10.5) | 12,763 (2.7) |
| Chronic obstructive<br>pulmonary diseases (%) | 932 (7.1) | 13,724 (3.0) | 650 (12.0) | 14,006 (3.0) |
| Dementia (%) | 140 (1.1) | 2,208 (0.5) | 88 (1.6) | 2,260 (0.5) |
| Diabetes mellitus (%) | 2,552 (19.5) | 30,595 (6.7) | 812 (15.0) | 32,335 (6.9) |
| Gout (%) | - | - | 258 (4.8) | 12,847 (2.8) |
| Heart failure (%) | 1,247 (9.5) | 8,712 (1.9) | 370 (6.8) | 9,589 (2.1) |
| Hypertensive diseases (%) | 7,807 (59.6) | 118,291 (25.7) | 2,890 (53.4) | 123,208 (26.3) |
| Immunodeficiencies (%) | 102 (0.8) | 1,664 (0.4) | 91 (1.7) | 1,675 (0.4) |
| Ischemic heart diseases (%) | 3,301 (25.2) | 42,250 (9.2) | 1,189 (22.0) | 44,362 (9.5) |
| Lipoprotein disorders (%) | 4,163 (31.8) | 58,824 (12.8) | 1,482 (27.4) | 61,505 (13.2) |
| Osteoarthritis (%) | 4,018 (30.7) | 75,353 (16.4) | 2,825 (52.2) | 76,546 (16.4) |
| Pulmonary heart disease (%) | 463 (3.5) | 6,002 (1.3) | 227 (4.2) | 6,238 (1.3) |
| Rheumatoid arthritis (%) | 258 (2.0) | 5,151 (1.1) | - | - |

SD = standard deviation.

**Table 2.**
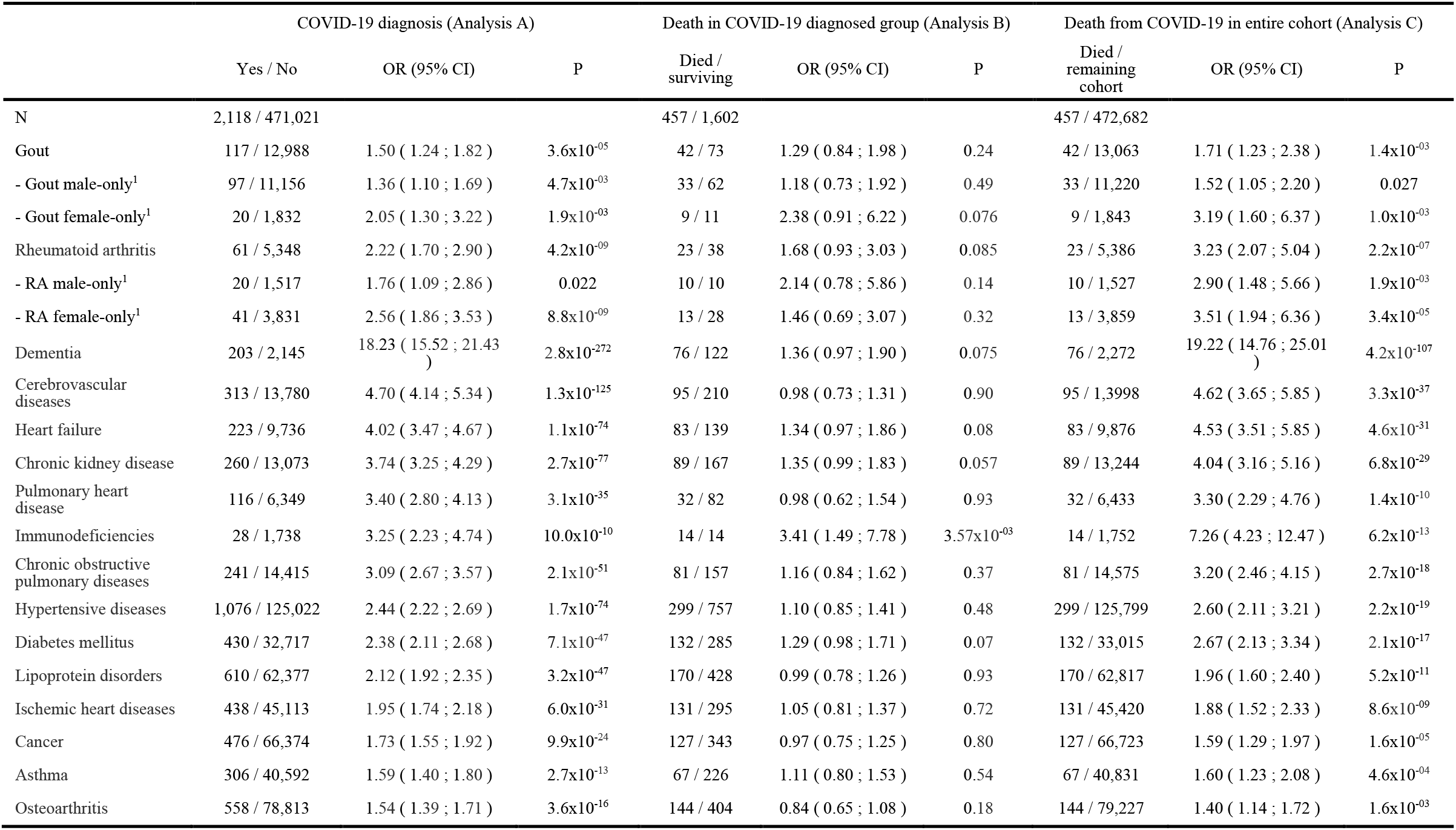
Logistic regression association analyses adjusted for current age, sex, ethnicity, Townsend deprivation index, BMI, and smoking-status (Model 1)

**Table 3.** Logistic regression association analyses adjusted by all other exposures listed in the table (Model 2).

|  | COVID-19 diagnosis (Analysis A) |  | Death in COVID-19 diagnosed group (Analysis B) |  | COVID-19-related death in entire cohort (Analysis C) |  |
| --- | --- | --- | --- | --- | --- | --- |
|  | OR (95% CI) | P | OR (95% CI) | P | OR (95% CI) | P |
| Male sex | 1.18 ( 1.08 ; 1.30 ) | 2.71x10<sup>-04</sup> | 1.44 ( 1.12 ; 1.85 ) | 4.00x10<sup>-03</sup> | 1.69 ( 1.38 ; 2.07 ) | 6.00x10<sup>-07</sup> |
| Age 60-69 years<sup>1</sup> | 0.54 ( 0.47 ; 0.61 ) | 6.96x10<sup>-21</sup> | 3.61 ( 1.99 ; 6.57 ) | 2.49x10<sup>-05</sup> | 1.97 ( 1.11 ; 3.49 ) | 0.021 |
| Age 70-74 years<sup>1</sup> | 0.45 ( 0.39 ; 0.53 ) | 2.50x10<sup>-26</sup> | 9.37 ( 5.18 ; 16.95 ) | 1.41x10<sup>-13</sup> | 3.70 ( 2.12 ; 6.46 ) | 4.09x10<sup>-06</sup> |
| Age > 74 years<sup>1</sup> | 0.60 ( 0.52 ; 0.69 ) | 1.33x10<sup>-13</sup> | 16.31 ( 9.20 ; 28.91 ) | 1.21x10<sup>-21</sup> | 7.30 ( 4.25 ; 12.53 ) | 5.60x10<sup>-13</sup> |
| Asian British<sup>2</sup> | 1.83 ( 1.48 ; 2.26 ) | 2.77x10<sup>-08</sup> | 0.65 ( 0.32 ; 1.33 ) | 0.24 | 1.10 ( 0.58 ; 2.09 ) | 0.77 |
| Black British<sup>2</sup> | 2.08 ( 1.68 ; 2.56 ) | 1.17x10<sup>-11</sup> | 1.56 ( 0.88 ; 2.76 ) | 0.13 | 2.72 ( 1.73 ; 4.28 ) | 1.50x10<sup>-05</sup> |
| Other ethnicities<sup>2</sup> | 1.54 ( 1.16 ; 2.04 ) | 3.10x10<sup>-03</sup> | 0.51 ( 0.19 ; 1.40 ) | 0.19 | 0.83 ( 0.34 ; 2.02 ) | 0.68 |
| Townsend deprivation index | 1.05 ( 1.04 ; 1.06 ) | 7.74x10<sup>-12</sup> | 1.04 ( 1.00 ; 1.07 ) | 0.044 | 1.07 ( 1.04 ; 1.10 ) | 7.44x10<sup>-06</sup> |
| BMI (per increase in kg/m<sup>2</sup>) | 1.02 ( 1.01 ; 1.03 ) | 1.27x10<sup>-05</sup> | 1.02 ( 1.00 ; 1.05 ) | 0.073 | 1.03 ( 1.01 ; 1.05 ) | 1.33x10<sup>-03</sup> |
| Current smoker<sup>3</sup> | 1.17 ( 1.06 ; 1.29 ) | 1.51x10<sup>-03</sup> | 1.05 ( 0.81 ; 1.37 ) | 0.70 | 1.27 ( 1.03 ; 1.58 ) | 0.027 |
| Ex-smoker<sup>3</sup> | 1.10 ( 0.95 ; 1.27 ) | 0.20 | 1.52 ( 1.04 ; 2.21 ) | 0.029 | 1.70 ( 1.25 ; 2.31 ) | 6.46x10<sup>-04</sup> |
| Gout | 1.01 ( 0.83 ; 1.24 ) | 0.91 | 1.26 ( 0.81 ; 1.95 ) | 0.31 | 1.18 ( 0.84 ; 1.65 ) | 0.35 |
| - Gout male-only | 1.01 ( 0.81 ; 1.26 ) | 0.91 | 1.20 ( 0.73 ; 1.99 ) | 0.47 | 1.15 ( 0.78 ; 1.69 ) | 0.47 |
| - Gout female-only | 0.96 ( 0.60 ; 1.54 ) | 0.86 | 2.16 ( 0.80 ; 5.86 ) | 0.13 | 1.71 ( 0.85 ; 3.46 ) | 0.13 |
| Rheumatoid arthritis | 1.34 ( 1.02 ; 1.77 ) | 0.038 | 1.63 ( 0.89 ; 2.96 ) | 0.11 | 1.89 ( 1.19 ; 3.02 ) | 7.24x10<sup>-03</sup> |
| - RA male-only | 1.00 ( 0.60 ; 1.66 ) | 0.99 | 2.33 ( 0.84 ; 6.52 ) | 0.11 | 1.50 ( 0.73 ; 3.07 ) | 0.27 |
| - RA female-only | 1.58 ( 1.13 ; 2.21 ) | 7.60x10<sup>-03</sup> | 1.41 ( 0.64 ; 3.09 ) | 0.39 | 1.96 ( 1.04 ; 3.67 ) | 0.037 |
| Dementia | 9.39 ( 7.89 ; 11.18 ) | 9.14x10<sup>-140</sup> | 1.33 ( 0.95 ; 1.88 ) | 0.10 | 10.18 ( 7.64 ; 13.58 ) | 2.67x10<sup>-56</sup> |
| Cerebrovascular diseases | 2.30 ( 2.00 ; 2.65 ) | 1.48x10<sup>-30</sup> | 0.93 ( 0.69 ; 1.26 ) | 0.64 | 2.08 ( 1.60 ; 2.71 ) | 5.04x10<sup>-08</sup> |
| Heart failure | 1.75 ( 1.47 ; 2.08 ) | 2.14x10<sup>-10</sup> | 1.27 ( 0.88 ; 1.84 ) | 0.20 | 2.05 ( 1.52 ; 2.76 ) | 2.57x10<sup>-06</sup> |
| Chronic kidney disease | 1.68 ( 1.44 ; 1.96 ) | 5.07x10<sup>-11</sup> | 1.23 ( 0.87 ; 1.72 ) | 0.24 | 1.69 ( 1.28 ; 2.23 ) | 1.83x10<sup>-04</sup> |
| Pulmonary heart disease | 1.93 ( 1.57 ; 2.37 ) | 4.12x10<sup>-10</sup> | 0.90 ( 0.56 ; 1.44 ) | 0.66 | 1.80 ( 1.22 ; 2.64 ) | 2.84x10<sup>-03</sup> |
| Immunodeficiencies | 1.99 ( 1.35 ; 2.93 ) | 5.31x10<sup>-04</sup> | 3.63 ( 1.58 ; 8.36 ) | 2.45x10<sup>-03</sup> | 4.58 ( 2.62 ; 8.01 ) | 9.77x10<sup>-08</sup> |
| Chronic obstructive pulmonary diseases | 1.64 ( 1.39 ; 1.93 ) | 3.09x10<sup>-09</sup> | 1.08 ( 0.76 ; 1.54 ) | 0.66 | 1.67 ( 1.25 ; 2.24 ) | 5.47x10<sup>-04</sup> |
| Hypertensive diseases | 1.57 ( 1.40 ; 1.75 ) | 4.07x10<sup>-15</sup> | 1.04 ( 0.78 ; 1.37 ) | 0.81 | 1.56 ( 1.23 ; 1.99 ) | 3.09x10<sup>-04</sup> |
| Diabetes mellitus | 1.36 ( 1.19 ; 1.54 ) | 3.37x10<sup>-06</sup> | 1.25 ( 0.93 ; 1.69 ) | 0.14 | 1.52 ( 1.19 ; 1.94 ) | 7.24x10<sup>-04</sup> |
| Lipoprotein disorders | 1.17 ( 1.03 ; 1.31 ) | 0.013 | 0.88 ( 0.67 ; 1.16 ) | 0.37 | 1.03 ( 0.82 ; 1.30 ) | 0.80 |
| Ischemic heart diseases | 0.92 ( 0.81 ; 1.05 ) | 0.23 | 0.92 ( 0.68 ; 1.24 ) | 0.57 | 0.85 ( 0.66 ; 1.10 ) | 0.21 |
| Cancer | 1.44 ( 1.29 ; 1.61 ) | 5.86x10<sup>-11</sup> | 1.00 ( 0.77 ; 1.30 ) | 0.98 | 1.29 ( 1.04 ; 1.61 ) | 0.020 |
| Asthma | 1.07 ( 0.93 ; 1.22 ) | 0.35 | 1.10 ( 0.78 ; 1.54 ) | 0.59 | 1.01 ( 0.76 ; 1.34 ) | 0.97 |
| Osteoarthritis | 1.08 ( 0.97 ; 1.21 ) | 0.16 | 0.78 ( 0.60 ; 1.01 ) | 0.059 | 0.93 ( 0.75 ; 1.16 ) | 0.54 |

<sup>1</sup> Reference group was < 60 years of age. <sup>2</sup> Reference group was white British; <sup>3</sup> Reference group was never smokers

### Associations with death upon diagnosis of COVID-19

When testing for association with death related to COVID-19 within the cohort with COVID-19 diagnosis (Analysis B), there was no evidence for association with gout or RA in either of Models 1 or 2 (Tables 2 and 3). For other diseases there was association with immunodeficiencies OR 3.4 [1.5 ; 7.8] (Model 1) and OR=3.6 [1.6 ; 8.4] (Model 2). Established risk factors for death from COVID-19, namely male sex and older age, were associated with death (OR 1.4 [1.1 ; 1.9] for males and, compared to <60 yrs, OR 3.6 [2.0 ; 6.6] for 60-69 yrs, OR 9.4 [5.2 ; 17.0] for 70-74 yrs, OR 16.3 [ 9.2 ; 28.9] for >74 yrs) (Table 3).

We then tested for association with death related to COVID-19 comparing to the entire UK Biobank cohort (Analysis C). Gout was associated with a 1.7-fold increase [95% CI: 1.2 ; 2.4] in COVID-19-related death under Model 1 but not in Model 2 (OR 1.2 [0.8 ; 1.7]). In contrast, RA associated with increased risk of death with COVID-19 in both models – OR 3.2 [2.1 ; 5.0] in Model 1 and OR 1.9 [1.2 ; 3.0] in Model 2. Given association of sex with prevalence of comorbidity in gout and RA (20-23) sex-specific analyses were performed. In RA the data were OR 2.9 [1.5 ; 5.7] for males and OR 3.5 [1.9 ; 6.4] for females in Model 1, and OR 1.5 [0.7 ; 3.1] and OR 2.0 [1.0 ; 3.7], respectively, in Model 2. In gout the data were OR 1.5 [1.1 ; 2.2] for males and OR 3.2 [1.6 ; 6.4] for females under Model 1, and OR 1.2 [0.8 ; 1.7] for males and OR 1.7 [0.9 ; 3.5] for females.

In Analysis C, we also assessed the 14 additional diseases for association with death from COVID-19 comparing to the entire UK Biobank cohort. Dementia, immunodeficiencies, chronic obstructive pulmonary diseases, cerebrovascular diseases, heart failure, pulmonary heart disease, chronic kidney disease, hypertensive diseases, diabetes mellitus, and cancer all associated with additional risk of death in Model 2 (Table 3), with dementia and immunodeficiencies having the strongest effects (OR 10.2 [7.6 ; 13.6] and 4.6 [2.6 ; 8.0], respectively). In Model 2, people of Black British ancestry had the highest risk of death (OR 2.7 [1.7 ; 4.3]), compared to people of White British ancestry, and there was positive association of death with BMI (OR = 1.03 [1.01 ; 1.05] per unit increase in BMI), and with an increased Townsend deprivation index score (OR=1.07 [1.04 ; 1.10]), consistent with a higher prevalence of seroprevalence of SARS-Cov-2 infection in people living in more deprived areas in the UK (24). Ex-smokers were at an increased risk of death with COVID-19 in model 2 (OR=1.7 [1.3 ; 2.3]) compared with never-smokers, consistent with the OpenSAFELY data from the UK (2), although directionality of association was different to the OpenSAFELY data for current smokers (OR=1.3 [1.0 ; 1.6]). Age group was also associated with death; OR 2.0 [1.1 ; 3.5] for 60-69 yrs, OR 3.7 [2.1 ; 6.5] 70 to 74 yrs, OR 7.3 [4.3 ; 12.5] for >74 yrs when compared to <60 yrs.

## Discussion

We identified RA as a risk factor for death related to COVID-19 in a multivariable-adjusted analysis of the UK Biobank cohort. Of clinical relevance, given implication of the type 1 interferon response in biologic therapy in RA (25), is involvement of a type 1 interferon-mediated immune response in people who die from COVID-19 (26), including in people with mutations in regulatory genes (27). It is important that the findings presented here are replicated in larger administrative datasets (eg. US-based National COVID Cohort Collaborative (www.ncats.nih.gov/n3c) and the UK OpenSAFELY cohort (2)). These datasets would allow for more stratification and use of additional models to fully explore factors including medications that might influence the observed association with RA. For example, the OpenSAFELY study included 962 individuals who died with COVID-19 who also had RA or SLE or psoriasis (2) – the number of people with RA in this group is likely to be at least 10-fold greater than in the UK Biobank data set used here. If the association we report here were replicated, investigation of the reasons for the relationship between RA and death from COVID-19 would improve understanding and potentially improve clinical management of COVID-19.

There are limitations to our analyses. Firstly, these analyses pertain to the population from which the UK Biobank was derived, predominantly the white European middle-aged ethnic group of the United Kingdom, and are not necessarily generalisable to other ethnic groups or other white European ethnic groups. There is also no available information on recovery status so there is the possibility of additional unidentified deaths in the COVID-19 diagnosed group in Analysis B. In addition to this COVID-19 outcomes will have been influenced over the time period of this study (March-August 2020) as clinical treatments evolved. General practice prescriptions were only available up until August 2019 and could not reliably be used to determine current medication usage. Thus the effect of anti-rheumatic treatments, particularly biologic disease-modifying antirheumatic drugs, could not be assessed in this study. Nor could the potential effect of disease activity in RA be assessed. Limited testing outside of the hospital settings means that the full extent of SARS-CoV-2 infection is not known in this population. Thus, it is not possible to accurately compare asymptomatic or mild COVID-19 to those with more severe disease. The UK Biobank dataset is also limited to those aged 49 to 86 as of 2020, a demographic with a higher infection fatality ratio (28). This will have contributed to the inflated infection fatality ratio in the UK Biobank cohort of 22%, well above general population estimates of 0.5 to 1.5% (e.g. (29)), in addition to a greater proportion of cases ascertained earlier in the pandemic by hospitalisation aligned with insufficient testing capability to detect community and mild cases (30,31). Therefore our findings cannot be generalised to those under 50 years of age. There is the potential in Analysis B for index event (collider) bias resulting from conditioning the sample set on COVID-19 diagnosis which would serve to bias towards the null (32). With respect to the lack of association of established risk factors for adverse COVID-19 outcomes (e.g. dementia) with death, the increased ascertainment of cases through hospitalisation earlier in the pandemic would also have contributed to bias towards the null in Analysis B. However these limitations were addressed using the entire cohort-based approach in Analysis C. We did not account for increased risk of death in RA for non-COVID-19-related causes which may have contributed to the OR of 1.80 (Table 3). However any inflation would have been countered by adjustment for multiple co-morbid conditions in Analysis C. A final limitation is that, while our method of ascertainment of gout in the UK Biobank has been validated (18), this is not the case for RA.

In summary, we found evidence for an effect of RA on the risk of death with COVID-19 independent of included comorbidities and known risk factors. This needs to be further explored in large datasets where a range of other factors can be investigated (e.g. RA therapies).

## Data Availability

All individual level data from which the results are derived are publicly-available from the UK Biobank.

## Acknowledgements

This research was conducted using the UK Biobank Resource under Application Number 12611. We sincerely thank all participants. The research was funded by the Health Research Council of New Zealand.

## Ethical approval

The UK Biobank resource was conducted under an ethical approval from the North West Multi-centre Research Ethics Committee (MREC) of the United Kingdom. The study complies with the Declaration of Helsinki and informed consent was obtained from all participants.

